# An inverse stage-shift model to estimate the excess mortality and health economic impact of delayed access to cancer services due to the COVID-19 pandemic

**DOI:** 10.1101/2020.05.30.20117630

**Authors:** Koen Degeling, Nancy N. Baxter, Jon Emery, Fanny Franchini, Peter Gibbs, G. Bruce Mann, Grant McArthur, Benjamin J. Solomon, Maarten J. IJzerman

## Abstract

**Background:** Decreased cancer incidence and reported changes to clinical management indicate that the COVID-19 pandemic will result in diagnostic and treatment delays for cancer patients. We aimed to develop a flexible model to estimate the impact of delayed diagnosis and treatment initiation on survival outcomes and healthcare costs based on a shift in the disease stage at treatment initiation.

**Methods:** The stage-shift model estimates population-level health economic outcomes by weighting disease stage-specific outcomes by the distribution of stages at treatment initiation, assuming delays lead to stage-progression. It allows for extrapolation of population-level survival data using parametric distributions to calculate the expected survival in life years. The model was demonstrated based on an analysis of the impact of 3 and 6-month delays for stage I breast cancer, colorectal cancer and lung cancer patients, and for T1 melanoma, based on Australian data. In the absence of patient-level data about time to stage progression, two approaches were explored to estimate the proportion of patients that would experience a stage shift following the delay: 1) based on the relation between time to treatment initiation and overall survival (breast, colorectal and lung cancer), and 2) based on the tumour growth rate (melanoma). The model is available on http://stage-shift.personex.nl/.

**Results:** A shift from stage I to stage II due to a 6-month delay is least likely for colorectal cancer patients, with an estimated proportion of 3% of the stage I patients diagnosed in 2020 progressing to stage II, resulting in 11 excess deaths after 5 years and a total of 96 life years lost over a 10-year time horizon. For breast and lung cancer, progression from stage I to stage II due to a 6-month delay were slightly higher at 5% (breast cancer) and 8% (lung cancer), resulting in 25 and 43 excess deaths after 5 years, and 239 and 373 life years lost over a 10-year time horizon, respectively. For melanoma, with 32% of T1 patients progressing to T2 disease following a 6-month delay, the model estimated 270 excess death after 5 years and 2584 life years lost over a 10-year time horizon.

**Conclusions:** Using a conservative 3-month delay in diagnosis and treatment initiation due to the COVID-19 pandemic, this study predicts nearly 90 excess deaths and $12 million excess healthcare costs in Australia over 5 years for the in 2020 diagnosed patients for 4 cancers. If the delays increase to 6 months, excess mortality and cost approach nearly 350 deaths and $46 million in Australia. More accurate data on stage of disease during and after the COVID-19 pandemic are critical to obtain more reliable estimates.

## 1. Introduction

National responses to the COVID-19 pandemic involved initiation of urgent responses to reduce the rate of new COVID-19 diagnoses and ensure intensive care capacity is sufficient to manage the potentially increased demand of acute hospital admissions. However, with social distancing measures being effective in controlling the immediate outbreak, additional concerns have emerged about delays in cancer diagnosis and treatment and the capacity of the health system to manage a backlog of patients who had not sought or received care during the acute phase of the pandemic.

Several countries have reported declining cancer incidences following COVID-19 and flagged this as an imminent policy concern, because it implies substantially delayed cancer diagnoses. The Netherlands Cancer Registry has presented weekly cancer incidence estimates since the outbreak of COVID-19, where the first case was reported on 27 February 2020. At the peak of the outbreak, cancer incidence was reduced by 40% on average, with the incidence rate for skin cancer decreasing 60% compared to historical data [1]. In addition, other changes have been reported, including an up to 66% reduction in chemotherapy administered across 8 hospitals in the United Kingdom [2]. Several professional societies have flagged these observations as requiring urgent action and encouraged interventions to restore the normal treatment throughput as much as possible [3].

This 2nd wave of pandemic consequences on the health service, which is not to be confused with a possible 2nd outbreak of COVID-19, is referred to as the impact of delayed access to care on health outcomes. These delays, as illustrated in Figure 1, may occur at different points throughout the care pathway [4]. In this terminology, there is an expected impact for patients due to diagnostic delays, caused by suspension of screening programs and delayed general practitioner (GP) visits or delayed access to diagnostic pathways. Additionally, there may be an impact of COVID-19 on care interruptions for patients already diagnosed and being treated for a chronic disease or cancer. For instance, care may be modified for immunocompromised patients at high risk of infection in the hospital setting by a delay or change in clinical management. Such changes include delaying surgery, a shift from intra-venously to orally administered systemic therapies, and modified radiotherapy delivery [5].

**Figure 1.**
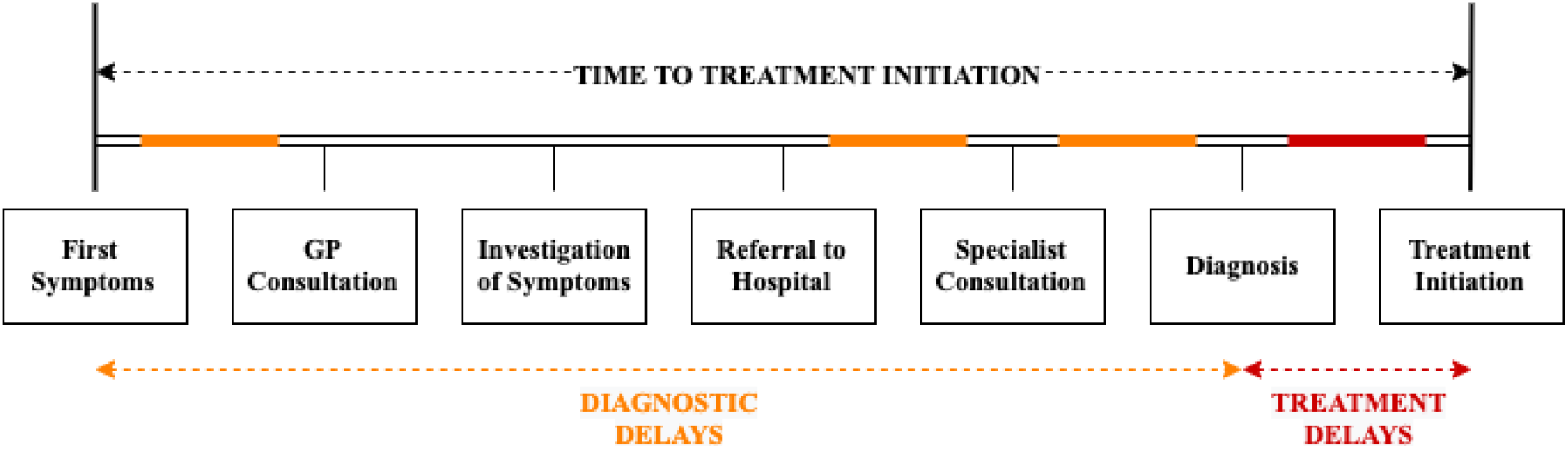
Based on Olesen et al. (2009) [5]: graphical illustration of different types of delays within the care pathway that may be induced by service disruptions due to the COVID-19 pandemic.

Two recent studies have estimated the health impact of COVID-19 on future cancer-specific health outcomes through estimates of excess mortality. Lai et al. (2020) used hospital admission data to estimate excess deaths of about 6,270 in the United Kingdom (UK) and 33,890 in the United States in addition to COVID-19 related mortality [2]. Sud et al. (2020) used observational studies to generate per-day hazard ratios of disease-stage specific cancer progression and assumed hypothetical delays of 3 and 6 months, estimating attributable deaths of 10,555 resulting from a 6 month delay [3].

Although both studies were well-designed and thought provoking, they were based on the impact of, for instance, short-term surgical delays or access to chemotherapy for which some evidence was available from the National Health Service (NHS) in the UK. However, delays due to the COVID-19 pandemic will not only occur in the hospital setting but also in the time to initial presentation and in the transition from primary to hospital care. Furthermore, delays will not have an equally negative effect on health outcomes for all cancer patients. While it is hard to estimate cancer specific excess mortality due to diagnostic and treatment delays, this study aims to address this problem through quantifying the impact of delays in time to treatment initiation (TTI) on cancer stage progression. Rather than assuming that TTI delays have a direct impact on health outcomes, this approach is based on the assumption that these delays only impact outcomes if they result in disease-stage progression, i.e. treatment being initialized at a more advanced disease stage [6].

Several studies have explored the relation between TTI and health outcomes. A systematic review concluded there is a relation between diagnostic delays and poorer outcomes for some cancers, including breast, colorectal, head and neck, testicular and melanoma [7]. Other studies also concluded a link between TTI and overall survival, with lung cancer and pancreas cancer being more heavily impacted by longer TTIs compared to breast cancer, for example [8]. Other recent studies concluded evidence of a relation between diagnostic delay and outcomes in head and neck cancers [9, 10]. For other cancers, like colorectal cancer, there is some evidence that delays to surgery of up to 30 days may not directly impact health outcomes [8, 11]. Despite varying levels of evidence of a relation between TTI and outcomes, studies conclude health systems should focus on the effectiveness of pathways after referral from primary care to shorten TTI [12]. Since responses to COVID-19 have led to restrictions for travel as well as imposed fears of patients to see health professionals, it is anticipated that delays in TTI will result in cancer progressing to a more advanced stage by the time of treatment initiation.

This paper presents a flexible stage-shift model that estimates long-term outcomes based on the distribution of disease stages at treatment initiation. It uses disease stage-specific incidence, survival and costing data to calculate expected survival in life years and excess healthcare costs for 4 cancers. The model, which is available in an online application, is versatile and flexible to include country specific data for any cancer type. The model is illustrated by estimating the impact of COVID-19 induced TTI delays on shifts from stage I to stage II for breast cancer, colorectal cancer and lung cancer, and from tumour stage 1 (T1) to T2 for melanoma. We will finally discuss limitations of the modelling approach.

## 2. Methods

The model developed to estimate the impact of delays in TTI is based on the distribution of disease stages at treatment initiation. It was implemented in an online tool using the shiny package [13] in R [14]: http://stage-shift.personex.nl. Although the tool allows data for any cancer type or country to be entered, Australian data for 4 cancers (breast cancer, colorectal cancer, lung cancer and melanoma) are included for demonstration. This section describes how the expected survival and healthcare costs are estimated and introduces the two approaches for calculating the stage shift resulting from delays in TTI. The R script used to perform the analysis is publicly available for download from the online tool. We refer to the *baseline scenario* as our best belief of the distribution of disease stages at treatment initiation in a scenario without delays, and the *stage shift scenario* as the distribution following delays.

### 2.1 Health impact in terms of expected life years

The expected survival in life years for a particular distribution of disease stages at treatment initiation is calculated based on the area under the combined survival curve. This combined survival curve is obtained by weighting stage-specific survival probabilities by the distribution of disease stage at treatment initiation (Figure 2). To allow for appropriate interpolation between data points and extrapolation beyond the time horizon of the data, a series of parametric distributions are fitted to the survival data. The survival function corresponding to the selected distribution is integrated to obtain the expected survival in life years for a specific scenario. The impact of a stage shift on the expected survival can be calculated as the difference between the survival outcomes for the *baseline scenario* compared to those of the *stage shift scenario*. Patient-level estimates of the expected life years are translated into population-level estimates based on the incidence.

**Figure 2.**
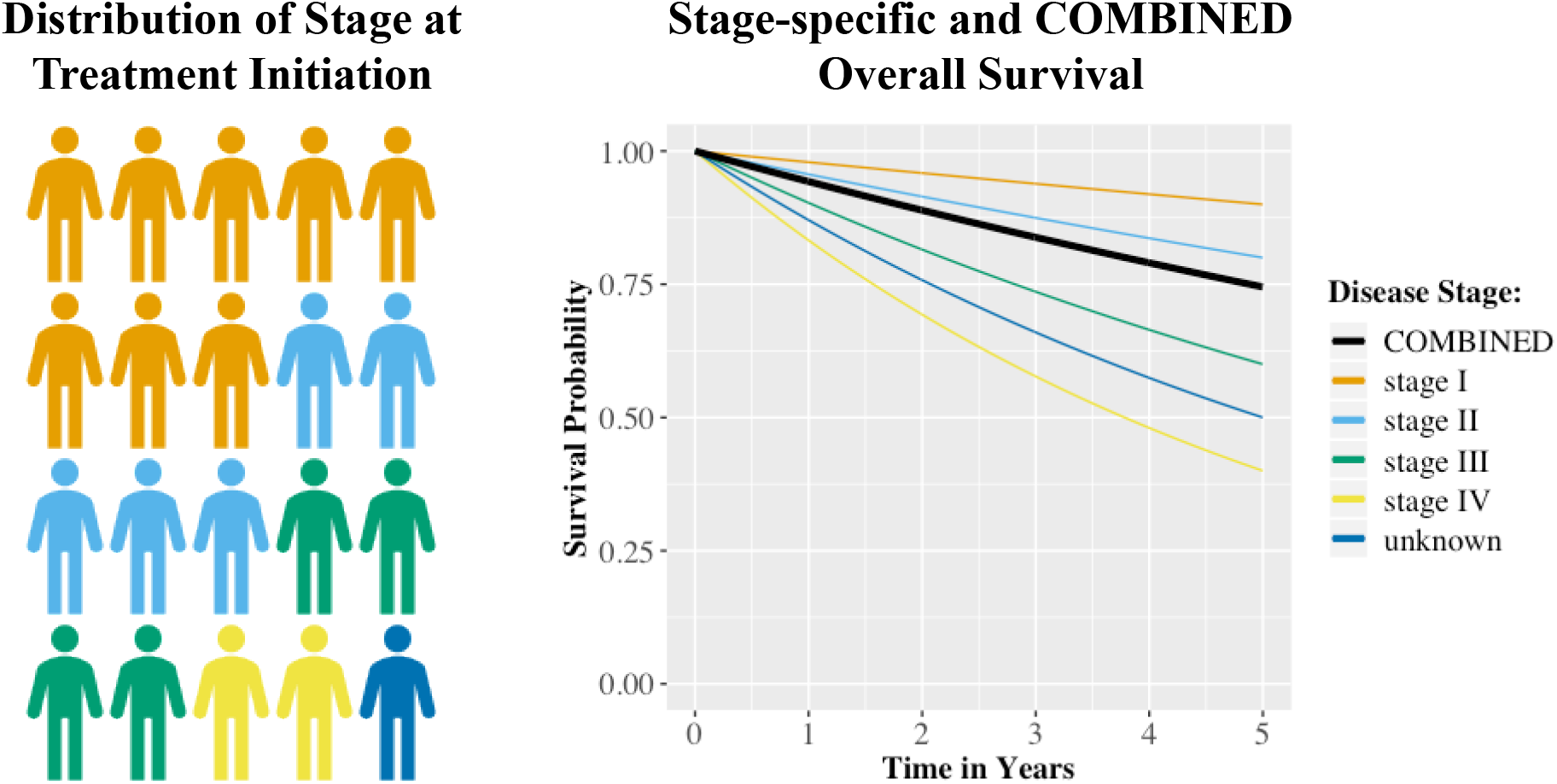
Graphical illustration of how the combined overall survival for a patient population is obtained by weighting stage-specific survival estimates by the stage distribution at treatment initiation.

The following distributions are fitted to the weighted survival curves using nonlinear least-squares regression of their parameters on logarithmic scale: exponential, Gamma, Gompertz, log-Normal, log-logistic, and Weibull. In absence of individual patient data, a parametric distribution cannot be selected using likelihood-based performance measures. Therefore, the tool allows for selection based on graphical and numerical inspection, including the survival probability point-estimates and the sum of the squared residuals.

To illustrate the framework, disease stage-specific (TNM stage) 1, 2, 3, 4, and 5-year *relative survival probabilities* were taken from the 2019 “Cancer data in Australia” report by the Australian Institute of Health and Welfare (AIHW) for breast cancer, colorectal cancer and lung cancer (Table 1) [15]. A different approach was taken to estimate the stage shift for melanoma patients (see Section 2.3.2), which required tumour (T) stage-specific survival data. Therefore, *overall survival probabilities* were estimated from the Melanoma Research Victoria (MRV) registry using data on 2,521 patients with a median follow up of 3.4 years. In the illustration, expected survival was estimated for time horizons of 5 and 10 years by extrapolating combined year-specific survival probabilities using Gompertz (breast cancer and melanoma) and Gamma (colorectal and lung cancer) distributions. Population-level estimates were based on the predicted 2020 incidence for Australia [15].

**Table 1.** Overview of the cancer type and stage-specific data used for estimating the impact in terms of survival and 5-year healthcare costs, with TNM classification and relative survival probabilities for breast, colorectal and lung cancer, and T-stage classification and overall survival probabilities for melanoma.

|  | Breast<br>Cancer | Colorectal<br>Cancer | Lung Cancer | Melanoma |
| --- | --- | --- | --- | --- |
| **Predicted incidence for 2020 (#)** | 19,998 | 16,634 | 13,092 | 15,691 |
| **Stage at Treatment Initiation (%)** |  |  |  |  |
| stage I / T1 | 43.0 | 22.1 | 11.7 | 61.8 |
| stage II / T2 | 34.7 | 24.3 | 6.5 | 13.6 |
| stage III / T3 | 12.1 | 23.6 | 11.2 | 10.2 |
| stage IV / T4 | 4.6 | 17.7 | 42.2 | 6.6 |
| stage unknown | 5.5 | 12.3 | 28.5 | 7.8 |
| **1-year survival (%)** |  |  |  |  |
| stage I / T1 | 100.0 | 99.3 | 90.8 | 99.2 |
| stage II / T2 | 99.9 | 96.4 | 69.8 | 98.0 |
| stage III / T3 | 98.1 | 93.5 | 57.8 | 97.2 |
| stage IV / T4 | 69.2 | 49.3 | 19.2 | 94.8 |
| stage unknown | 89.5 | 75.9 | 43.6 | 89.9 |
| **2-year survival (%)** |  |  |  |  |
| stage I / T1 | 100.0 | 99.1 | 82.7 | 97.9 |
| stage II / T2 | 98.8 | 94.2 | 52.4 | 94.5 |
| stage III / T3 | 93.4 | 85.5 | 32.9 | 89.5 |
| stage IV / T4 | 57.6 | 32.0 | 8.3 | 65.0 |
| stage unknown | 83.1 | 65.8 | 27.1 | 80.7 |
| **3-year survival (%)** |  |  |  |  |
| stage I / T1 | 100.0 | 98.8 | 76.1 | 96.9 |
| stage II / T2 | 97.5 | 92.9 | 42.0 | 89.6 |
| stage III / T3 | 88.3 | 79.1 | 25.2 | 83.9 |
| stage IV / T4 | 47.1 | 22.8 | 5.3 | 75.5 |
| stage unknown | 76.4 | 61.4 | 20.4 | 76.3 |
| **4-year survival (%)** |  |  |  |  |
| stage I / T1 | 100.0 | 99.0 | 71.4 | 94.4 |
| stage II / T2 | 95.7 | 90.8 | 35.6 | 85.8 |
| stage III / T3 | 84.2 | 74.3 | 20.1 | 78.8 |
| stage IV / T4 | 38.7 | 16.8 | 3.9 | 67.5 |
| stage unknown | 69.9 | 58.7 | 16.1 | 74.6 |
| **5-year survival (%)** |  |  |  |  |
| stage I / T1 | 100.0 | 98.6 | 67.7 | 91.7 |
| stage II / T2 | 94.6 | 88.6 | 32.3 | 83.0 |
| stage III / T3 | 80.6 | 71.3 | 17.1 | 73.2 |
| stage IV / T4 | 32.0 | 13.4 | 3.2 | 61.6 |
| stage unknown | 65.0 | 56.9 | 14.4 | 73.7 |
| **Healthcare costs over 5 years** |  |  |  |  |
| stage I / T1 | \$50,699 | \$51,531 | \$28,532 | \$13,981 |
| stage II / T2 | \$67,069 | \$66,504 | \$28,532 | \$25,715 |
| stage III / T3 | \$98,632 | \$121,558 | \$67,065 | \$30,644 |
| stage IV / T4 | \$105,934 | \$120,015 | \$79,045 | \$38,216 |
| stage unknown | \$100,860 | \$121,077 | \$70,678 | \$32,960 |

### 2.2 Economic impact in terms of healthcare costs

The expected healthcare costs are calculated as the stage-specific costs weighted according to the distribution of disease stage at treatment initiation. In the illustration, the impact of stage shifts on healthcare costs in Australia was estimated up to 5 years after diagnosis from a public healthcare payer perspective. Goldsbury et al. (2018) used data of 7,624 participants with a cancer diagnosis registered in the New South Wales Cancer Registry between 2006 and 2010, matched to 22,661 cancer-free controls, to determine excess healthcare costs of the 10 most common cancers in Australia [16]. Healthcare costs considered included those associated with inpatient hospital episodes, emergency department presentations, medicines captured in the Pharmaceutical Benefits Scheme (PBS) and medical services captured in the Medicare Benefits Schedule (MBS).

Goldsbury et al. did not publish disease stage-specific costs that were needed to estimate the impact of stage shifts. Therefore, stage-specific costs were approximated for each cancer separately in three steps (see Appendix A for details). First, the total expected treatment costs over the five years after diagnosis were determined based on the year-specific costs presented by Goldsbury et al. and the combined survival probabilities over all disease stages weighted using the stage distribution according to the AIHW data or MRV data. Second, the distribution of costs between disease stages were approximated using other costing studies [17–20]. Finally, based on the total costs over all stages and the distribution of costs between stages, the stage-specific costs were determined (Table 1). For lung cancer, none of the identified studies distinguished between healthcare costs for stage I and stage II disease, so these were assumed to be equivalent and based on the most recent study [19, 21–23]. Because of a lack of evidence regarding the cost of care for cancer patients with an unknown disease stage at diagnosis, stage-specific costs of the other stages were weighted based on coefficients estimated in a least-squares regression weighting the stage-specific survival probabilities to match those of the unknown stage category. All costs were indexed to 2020 Australian dollars ($) using the Australian Health Index [24].

### 2.3 Distribution of disease stage at treatment initiation

The framework considers up to 5 disease stage categories to define the *baseline* and *stage shift scenario*. Although these are classified as stage I, stage II, stage III, stage IV and unknown stage (TNM stages) in the online tool, they can also be used for other classifications, for example according to T stage as in the illustration for melanoma. The illustration here used data on the distribution of disease stage published by the AIHW [15] to define the *baseline scenario* for breast, colorectal and lung cancer, and data published by Cancer Council Victoria for the distribution of T stage for melanoma [25].

In the absence of empirical evidence on the distribution of disease stage at treatment initiation during the COVID-19 pandemic, 2 approaches were explored and compared to approximate this distribution through estimating time to stage progression (TTSP). The first approach was based on the relation between TTI and survival (Section 2.3.1), whereas the second approach was based on the tumour growth rate (Section 2.3.2). The first approach was applied for breast, colorectal and lung cancer, and the second approach was applied for melanoma. To be conservative in these exploratory analyses and reduce the complexity of the illustration, only shifts from stage I to stage II were modelled for breast, colorectal and lung cancer, and from stage T1 to T2 for melanoma. Two realistic delay periods were considered: 3 months and 6 months.

#### 2.3.1 Stage progression based on the relation between TTI and survival

The *treatment delay approach* was based on the relation between TTI and survival, estimating how long a delay in TTI needs to be such that the mortality rate for stage I patients matches the rate for stage II patients. For each cancer type separately, stage-specific excess mortality rates, denoted here by 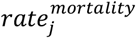, were calculated from 5-year relative survival probabilities reported by the AIHW [15]. Here, *j* = *stage I, stage II*, but this also generalizes to more stage categories. The required hazard ratio 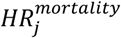 for a 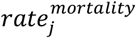 to match the rate of the subsequent stage *j* + 1 was calculated as the ratio between the two rates:

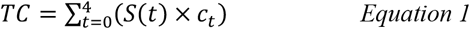

Subsequently, using a stage-specific hazard ratio of a delay in TTI on overall survival 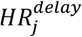 and corresponding delay in 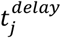 reported in literature, the expected delay 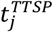 required for a stage shift was determined by assuming a linear relationship between the delays and hazard ratios on logarithmic scale (Equation 2). This resembles a continuous variable in a Cox-proportional hazards model, assuming an exponential relation between a delay in TTI and overall survival.

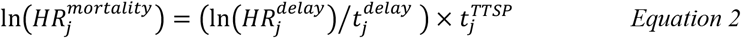

Since 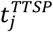 represents the expected value of the delay in TTI required for a stage shift to occur, the corresponding event rate for TTSP can be calculated as follows:

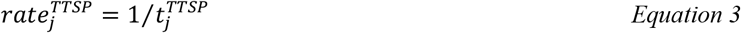

This event rate can be used to define an exponential distribution to calculate the probability that a patient will progress given a certain delay. Since the 100% relative 5-year survival for stage I breast cancer in Australia reported by the AIHW does not allow this approach to be applied, because it suggests an excess mortality rate of zero, 5-year relative survival was assumed to be 99.9% in calculating 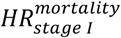 for breast cancer. Hazard ratios for 1-week TTI delays on overall survival for stage I patients extracted from a large population study were 1.018 (breast cancer), 1.005 (colorectal cancer) and 1.032 (lung cancer) [8], yielding TTSP estimates of 4.3 years, 8.3 and 2.9 years, respectively. This study used data from the National Cancer Database (NCDB) (breast cancer: n = 1,368,024; colorectal cancer: n = 662,094; lung cancer: n = 363,863) to estimate the impact of time between clinical or histologically confirmed diagnosis and the start of any cancer-directed treatment on survival, considering time beyond 6 weeks as delays. The TTSP estimates were used to calculate the proportion of patients that would experience a stage progression in each month in which services were disrupted, assuming incident cases are diagnosed evenly throughout the year. These proportions were then applied to the *baseline scenario* to define the *stage shift scenario* for the 3 and 6 month delays separately.

#### 2.3.2 Stage progression based on tumour growth

The *tumour growth approach* estimated the proportion of patients that will progress to a more advanced stage based on the tumour growth rate. This approach was illustrated for melanoma, where the T stage is directly linked to the thickness of the tumour (Breslow thickness) and evidence on the monthly increase of tumour thickness is available. Liu et al (2006) have reported an average growth in tumour thickness of 0.08 millimetres per month for tumours that are up to 1 millimetre in thickness based on individual patient reports of times from first change in a lesion to a pathological diagnosis [26]. Based on this growth rate and by assuming a Uniform distribution for tumour thickness for T1 tumours at treatment initiation, it was simulated how many patients would have progression to a T2 tumour (i.e., with a thickness between 1 and 2 millimetres) following a 3 or 6-month delay. This Uniform distribution was defined with a minimum and maximum of 0.25 and 1 millimetre, respectively, based on inspection of empirical T1 tumour thickness data from the MRV registry (n = 710). The resulting simulated mean thickness without a delay of 0.63 millimetres matched the observed mean of 0.64 millimetres well. The estimated proportions were applied to the *baseline scenario* to define the *stage shift scenarios*.

## 3. Results

Table 2 presents an overview of the results of the analyses performed using the stage-shift framework for a 3 and 6-month delay in TTI for the 2020 Australian incident breast cancer, colorectal cancer, lung cancer and melanoma patient populations.

**Table 2.**
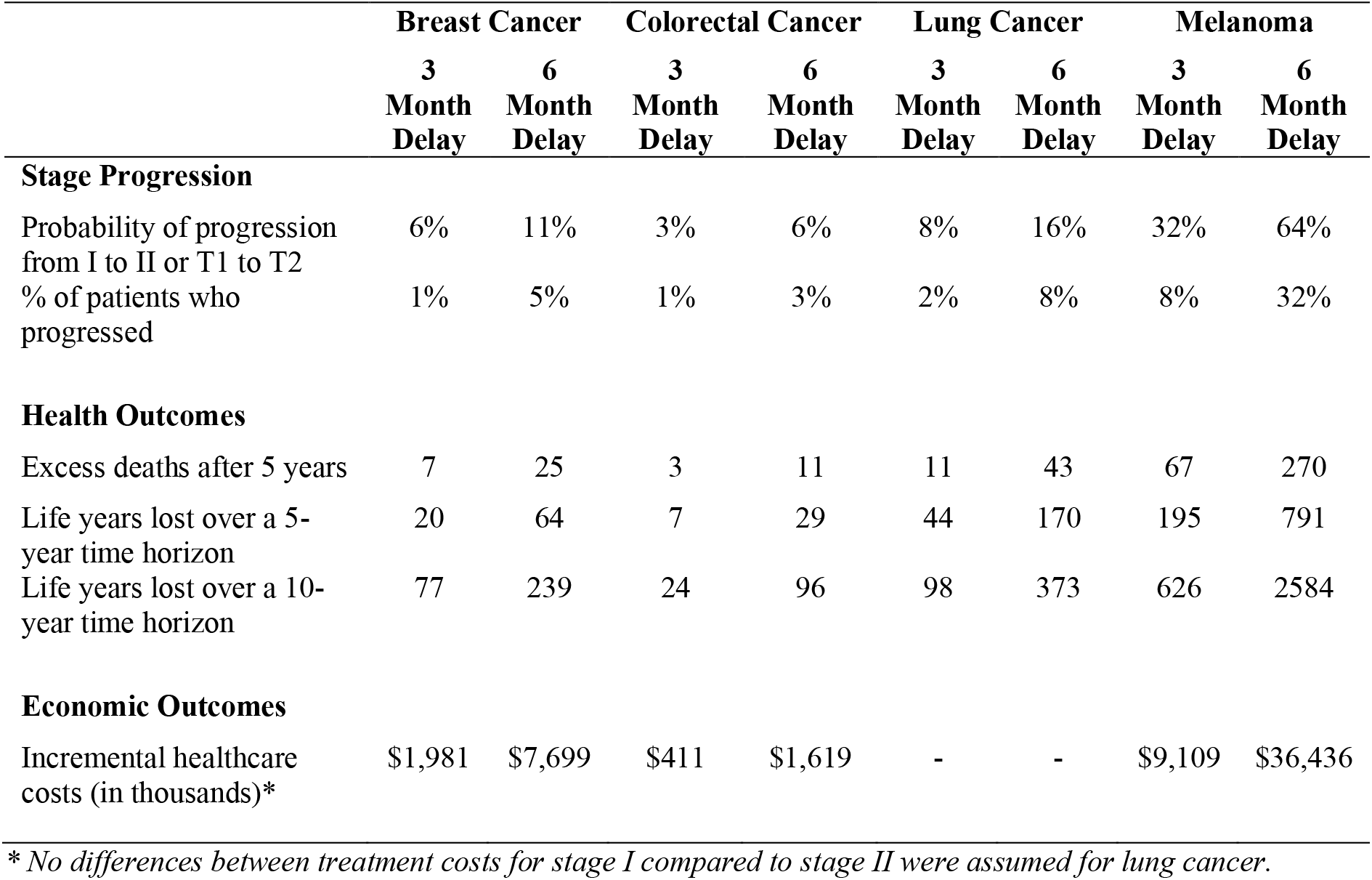
Results of the exploratory analyses for the 2020 Australian incident populations, only considering shifts from stage I to stage II (breast cancer, colorectal cancer and lung cancer) and stage T1 to stage T2 (melanoma).

|  | Breast Cancer |  | Colorectal Cancer |  | Lung Cancer |  | Melanoma |  |
| --- | --- | --- | --- | --- | --- | --- | --- | --- |
|  | 3<br>Month<br>Delay | 6<br>Month<br>Delay | 3<br>Month<br>Delay | 6<br>Month<br>Delay | 3<br>Month<br>Delay | 6<br>Month<br>Delay | 3<br>Month<br>Delay | 6<br>Month<br>Delay |
| **Stage Progression** |  |  |  |  |  |  |  |  |
| Probability of progression from I to II or T1 to T2 | 6% | 11% | 3% | 6% | 8% | 16% | 32% | 64% |
| % of patients who progressed | 1% | 5% | 1% | 3% | 2% | 8% | 8% | 32% |
| **Health Outcomes** |  |  |  |  |  |  |  |  |
| Excess deaths after 5 years | 7 | 25 | 3 | 11 | 11 | 43 | 67 | 270 |
| Life years lost over a 5-year time horizon | 20 | 64 | 7 | 29 | 44 | 170 | 195 | 791 |
| Life years lost over a 10-year time horizon | 77 | 239 | 24 | 96 | 98 | 373 | 626 | 2584 |
| **Economic Outcomes** |  |  |  |  |  |  |  |  |
| Incremental healthcare costs (in thousands)* | \$1,981 | \$7,699 | \$411 | \$1,619 | - | - | \$9,109 | \$36,436 |

\* No differences between treatment costs for stage I compared to stage II were assumed for lung cancer.

For a delay in TTI of 3 months for stage I breast cancer patients, the *treatment delay approach* estimated a 6% probability that these patients would have progressed to stage II during this period of disruption. For a 6-month delay, this probability was estimated to be 11%. Because not all stage I patients in the 2020 population would have been diagnosed in the months where COVID-19 restrictions were imposed, the adjusted proportion of stage I patients experiencing progression were 1% and 5%, for a 3 and 6-month delay respectively. The stage-shift model then estimated this would result in 7 or 25 excess deaths after 5 years for the 2020 breast cancer population, respectively. Over a 5 and 10-year time horizon, the estimated expected loss in life years for the 2020 population was 20 and 77 life years if a 3-month delay was observed, and 64 and 239 life years for a 6-month delay. Following the stage-shift due to a 3-month delay, healthcare costs for the incident 2020 breast cancer population were estimated to increase with $2.0 million, whereas a $7.7 million increase was estimated for a 6-month delay.

Because the TTSP estimated using the *treatment delay approach* for colorectal cancer exceeded the TTSP for breast cancer, the probability of stage progression was lower compared to breast cancer at 3% for a 3-month delay and 6% for a 6-month delay, which translated to proportions of the 2020 incident stage I population of 1% and 3%, respectively. The number of excess deaths after 5 years were estimated to be 3 and 11, for a 3 and 6-month delay respectively. Over a 5 and 10-year time horizon, the loss in expected survival was 7 and 24 life years for a 3-month delay, and 29 and 96 life years for a 6-month delay. The increment in healthcare costs for a 3-month delay in the 2020 diagnosed population was estimated at $411,000, with an estimated increase of $1.6 million for a 6-month delay.

For lung cancer, the probability of progression during a 3-month delay was estimated to be 8% according to the *treatment delay approach*, and 16% for a 6-month delay. Consequently, the proportion of stage I lung cancer patients from the 2020 population that would progress during the delay was 2% and 8%, for a 3 and 6-month delay respectively. Given the more substantial difference in overall survival between stage I and stage II for lung cancer compared to breast and colorectal cancer, this translated into estimated excess deaths of 11 and 43 after 5 years, respectively. In terms of expected survival in life years, a 3-month delay was estimated to result in a loss of 44 and 98 life years, and a 6-month delay to result in a loss of 170 and 373 life years, over a 5 and 10-year time horizon respectively. No differences in healthcare costs for treating the incident 2020 lung cancer population following a stage shift from stage I to stage II were found, because treatment costs for these two stages were estimated to be equal (Table 1).

Melanoma was the only tumour stream for which stage progression following a delay in TTI was estimated using the *tumour growth approach*, which contributed to the highest impact across all considered cancer types. The probabilities of progressing due to a 3 or 6-month delay were 32% and 64%, respectively, which was estimated to result in 8% or 32% of the 2020 T1 melanoma patients progressing to T2 disease. At 67 following a 3-month delay and 270 following a 6-month delay, the number of excess deaths after 5 years was the highest for melanoma. This translated into 195 or 626 life years lost for a 3-month delay, and 791 or 2584 life years lost for a 6-month delay, over a 5 and 10-year time horizon respectively. Based on the stage shift, healthcare costs for treating the 2020 diagnosed T1 melanoma population were estimated to increase with $9.1 million following a 3-month delay or $36.4 million following a 6-month delay.

## 4. Discussion

Here we present an early modelling study to estimate excess mortality due to delays in TTI as a result of COVID-19. For the 4 cancers included, exploratory analyses estimated that the excess mortality projected across Australia can be up to 90 patients after 5 years if delays up to 3 months are considered, which increases to nearly 350 deaths if 6-month delays are experienced. In addition, excess healthcare costs can be up to $12 million over 5 years for a 3-month delay, increasing to $46 million for a 6-month delay. Compared to the limited number of deaths from COVID-19 in Australia (102 deaths on 26 May 2020), the excess mortality therefore causes an imminent policy concern. Particularly if new cancer diagnoses continue to remain low compared to baseline and further delays occur if hospital and surgical capacity is affected, the long-term effect of COVID-19 on cancer health outcomes should not be neglected.

This study is not the first to flag the concerns of increasing diagnostic and treatment delays. A rapid decline in cancer incidence in the Netherlands has led to immediate awareness campaigns that encouraged people to see their GP if they experienced suspicious symptoms [1]. Earlier studies have also flagged the collateral damage occurring from declining cancer incidence and reductions in delivery of multi-disciplinary cancer services in the UK [2, 3].

This study uses a stage-shift model to illustrate the impact of delays, which was illustrated for 4 cancers. While the approach is flexible and easy to implement, one criticism of the approach is that stage at treatment initiation is assumed to be an ordinal variable for which survival and costs were extrapolated. However, there is significant heterogeneity within each stage between patients with poorer and better prognoses (e.g., tumour size of 4 mm compared to 17mm in breast cancer). Nevertheless, compared to previous modelling approaches, a strength of the current model is that it does not assume that the delays will affect all patients, but only those whose disease progresses to a subsequent, more advanced stage. This is an essential feature, as current observational studies and systematic reviews are not conclusive about the relation between diagnostic delays and survival and significant heterogeneity is expected between cancers and between individual patients [7].

Because patient-level data about the disease stage at treatment initiation during the COVID-19 pandemic is not yet available, we estimated this distribution based on the delay in TTI. For this estimate we extensively searched the literature for observational studies investigating the relation between treatment delay, stage progression and survival [8, 27–29], as well as mathematical models predicting stage progression from tumour growth [30, 31]. The likelihood of stage shifts according to the *treatment delay approach* strongly depends on the assumed hazard ratios for the impact of delays in TTI on survival. In the illustration for stage I breast cancer, colorectal cancer and lung cancer, these hazard ratios were extracted from recent study that used a large patient sample from the NCDB to obtain estimates for multiple cancer types according to a consistent analysis framework. Although these estimates from a single study improve comparability between cancer types, other studies have also estimated the impact of delays in TTI on survival. For breast cancer, Bleicher et al. (2016) used data from the NCDB (n = 115,790) and Surveillance, Epidemiology and End Results (SEER) database (n = 945,441) to estimate hazard ratios of 1.16 (NCDB) and 1.13 (SEER) for every 30-day increase in time between diagnosis and surgery for stage I [28]. These estimates are higher compared to the hazard ratio of 1.018 for a 7-day delay used here, which translates to a 30-day hazard ratio of 1.08. For colorectal cancer, Lee et al. (2019) used data on 39,900 patients from the Taiwan Cancer Registry Database to estimate a hazard ratio of 1.41 for TTI between 31 and 150 days compared to TTI < 31 days for stage I, and a hazard ratio of 2.66 for TTI beyond 151 days [29], which are substantially higher than the hazard ratio used in the illustration. Regarding lung cancer, Samson et al. (2015) used data on 39,995 stage I non-small cell lung cancer patients from the NCDB to estimate an hazard ratio of 1.004 for every week between diagnosis and surgery beyond 8 weeks [32], which is lower than the hazard ratio of 1.032 used here.

While different assumptions had to be made, we aimed to provide conservative but realistic estimates based on the Australian data and by analysing a realistic 3-month and a possible 6-month delay. First, although the impact of stage progressions from stage II to stage III or from stage III to stage IV are likely to be associated with significant excess mortality and cost, we only considered shifts from stage I to stage II disease for breast, colorectal and lung cancer, and shifts from T1 to T2 for melanoma. These shifts have a relatively limited impact on expected survival and healthcare expenditure. Second, because the illustration focusses on stage I and T1 cancers, only a proportion of the complete patient populations was considered to experience delays and be at risk of disease progression: 62% for melanoma, 43% for breast cancer, 22% for colorectal cancer and 12% for lung cancer (Table 1). Since diagnostic and treatment delays are likely to occur throughout all stages of disease, the estimates in our illustration are relatively conservative, because those patients would also be at risk of disease progression. Diversity in the proportion of stage I or T1 patients between the 4 cancers relative to their absolute incidence also explains differences in excess mortality, with the impact in breast cancer and melanoma being more severe. If we had modelled additional stage shifts from stage III to stage IV, for example, the excess mortality in lung and colorectal cancer would have been substantially higher. Third and finally, we only model incident cases, i.e. patients diagnosed in 2020. But delays in treatment for prevalent patients, such as surgery, have also been reported. Furthermore, it is likely that prevalent cancer patients at high-risk for COVID-19 will experience additional service disruptions, such as adjusted therapy intensity. Obviously, when considering the impact of treatment delays and modifications for prevalent patients, real-world excess mortality is expected to be substantially higher.

In conclusion, we have developed a flexible stage-shift model allowing estimation of the long-term health outcomes and economic impact of delays in access to care due to the COVID-19 pandemic. Initial analyses have estimated an excess mortality of nearly 90 deaths and excess costs of more than $12 million based on a realistic 3-month delay. However, the analyses also demonstrate that hundreds of life years may be lost over the following years due to delays in TTI if delays increase to 6 months. Although the results are preliminary, they are indicative of the potential impact of service disruptions due COVID-19 on cancer outcomes in the years to come and can create awareness and provide incentive for restoring health service throughput levels to normal as soon as possible.

## Data Availability

The framework has been made publicly available in an online tool: http://stage-shift.personex.nl. All data used to illustrate the framework are included in this publication. The R scripts used to perform the exploratory analyses within the illustration is available from the online tool.

## Acknowledgements

The Melanoma Research Victoria registry, from which data has been used in this study, was supported by the Victorian Government through the Victorian Cancer Agency Translational Research Program.

## Appendix A details on approximation of stage-specific healthcare costs

The modelling framework uses disease stage-specific healthcare costs to estimate the overall economic impact of a shift in disease stage at treatment initiation. It does so by comparing the expected costs on patient or population level according to the baseline scenario, which represents our best belief about how the distribution of disease stages at treatment initiation without a stage shift, to those of the stage shift scenario, which represents the distribution of disease stages following the stage shift. For each scenario, the expected costs are obtained by weighting the stage-specific costs by the respective distribution of disease stages at treatment initiation. This appendix details how stage-specific healthcare costs were obtained for the illustration of the modelling framework.

The illustration included estimates of the economic impact of stage shifts following COVID-19 induced delays in the time to treatment initiation for breast cancer, colorectal cancer, lung cancer and melanoma from an Australian healthcare payer perspective. The study by Goldsbury et al. (2018) [1] was preferred as a single source of evidence for Australian costing data for all considered cancer, because costing estimates from different costing studies are often incomparable due to differences in healthcare system context or study design. However, Goldsbury et al. do not present stage-specific costing data, which are required as inputs for the modelling. Hence, stage-specific cost estimates were obtained by synthesizing data presented by Goldsbury et al. with information of the distribution of costs over disease stages from other publications in three steps:

1. the total expected costs over the 5 years after diagnosis were estimated based on the year-specific costs presented by Goldsbury et al. and the combined survival probabilities over all disease stages;
2. the distribution of costs between disease stages were approximated using information from other studies;
3. the stage-specific costs were estimated so that the combined costs weighted according to the distribution of disease stages at treatment initiation in the baseline scenario result in the total expected costs (Step 1), while matching the distribution of costs between stages (Step 2).

### Step 1: estimating the total costs over the 5 years after diagnosis

Goldsbury et al. present year-specific excess costs for the 5 years after diagnosis per patient that received care in that year. To calculate the total expected costs over this period, it would be wrong to sum these year-specific costs, because not every patient will be alive throughout the 5 years after diagnosis. Hence, the expected value of the total 5-year healthcare costs *TC* is calculated by taking into consideration the survival probabilities, using the following formula:

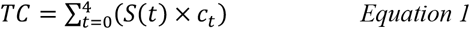

where *t* represents the year after diagnosis, *S*(*t*) is the survival probability, and *c*_*t*_ represents the year-specific costs per patient who is receiving care. Note that *t* = 0 represents the first year after diagnosis, where all patients are assumed to receive care, i.e. *S*(0) = 1. The stage-specific survival probabilities are reported in the main manuscript and were weighted according to the baseline scenario to obtain the combined survival *S*(*t*) for each cancer separately. Note that for melanoma this was based on tumour (T) stage rather than overall disease stage (TNM). Based on these data and the data from Goldsbury et al, the total expected excess healthcare costs over the first 5 years after diagnosis presented in Table A1 were calculated.

**Table A1.** Year-specific costs per patient receiving care presented in Goldsbury et al. (2018) and expected total excess healthcare costs over the first 5 years after diagnosis in 2013 Australian dollars ($).

| Cancer Type | Year-specific costs from Goldsbury et al. (2018) |  |  |  |  | Expected Total Costs |
| --- | --- | --- | --- | --- | --- | --- |
|  | Year 0 - 1 | Year 1 - 2 | Year 2 - 3 | Year 3 - 4 | Year 4 - 5 |  |
| Breast Cancer | \$36,948 | \$7,619 | \$3,038 | \$3,132 | \$2,473 | \$52,526 |
| Colorectal Cancer | \$48,570 | \$10,926 | \$7,381 | \$6,509 | \$4,525 | \$71,829 |
| Lung Cancer | \$28,059 | \$17,863 | \$12,903 | \$6,563 | \$4,021 | \$51,435 |
| Melanoma | \$7,110 | \$2,260 | \$2,792 | \$2,506 | \$1,784 | \$15,828 |

### Step 2: defining the distribution of costs between disease stages

To approximate the stage-specific costs based on the reported overall costs by Goldsbury et al, the relation between costs across the different stages were based on other studies (Table A2). For breast cancer, a global systematic review reporting stage-specific costs was used [2], since this was the only study to distinguish between stage I and stage II disease. An Australian costing study was used for colorectal cancer [3]. No studies that distinguish between stage I and stage II lung cancer were found, so a recent study from the United States (US) was used [4]. A US study was also used for melanoma, assuming the relation between disease stage-specific costs for T-stage to be equivalent to the distribution of costs according to TNM classification [5]. These stage-specific costs were used solely to define the relationship between healthcare costs for different stages.

**Table A2.** Disease stage-specific costs extracted from literature (currency not of relevance).

| Cancer Type | Stage I | Stage II | Stage III | Stage IV | Reference |
| --- | --- | --- | --- | --- | --- |
| Breast Cancer | 29,724 | 39,322 | 57,827 | 62,108 | Sun et al. (2019) |
| Colorectal Cancer | 34,952 | 45,108 | 82,449 | 81,403 | Ananda et al. (2016) |
| Lung Cancer | 2,636 | 2,636 | 6,196 | 7,303 | Sheehan et al. (2019) |
| Melanoma | 14,499 | 26,667 | 31,778 | 39,631 | Seidler et al. (2010) |

Since the studies did not report on healthcare costs for patient with an unknown stage, this needed to be estimated, which was done by assuming the “stage unknown” category is a mix of stage III and stage IV patients for breast cancer, colorectal cancer and lung cancer, and a potential mix of T1, T2, T3 and T4 patients for melanoma. The weight of each stage was determined through a least-squares regression matching the weighted stage-specific survival to the survival for the “stage unknown” category. The results of this analysis are presented in Table A3.

**Table A3.** Contributions of disease stages to the “unknown stage” category used to define the relation between costs for this category relative to the other categories.

| Cancer Type | Stage I | Stage II | Stage III | Stage IV |
| --- | --- | --- | --- | --- |
| Breast Cancer | - | - | 69% | 31% |
| Colorectal Cancer | - | - | 69% | 31% |
| Lung Cancer | - | - | 70% | 30% |
| Melanoma | 22% | 0% | 0% | 78% |

### Step 3: estimating the stage-specific healthcare costs

The final step to approximate the stage-specific costs was to estimate them in such a way that the combined costs weighted according to the distribution of disease stages at treatment initiation in the baseline scenario were equivalent to the expected total costs (Step 1), while matching the distribution of costs between stages (Step 2). This can be done, for example, through the “Goal Seek” function in Microsoft Excel, a least-squares regression or other optimization algorithm. More specifically, by setting the costs for stage I, for example, the costs for the other stages were defined through the relations defined in Step 2, which then defined the combined costs according to the distribution of stages. The value for the costs of stage I was optimized such that the weighted combined costs matched the expected total costs estimated in Step 1. The results of this step are presented in Table A4.

**Table A4.** Approximated stage-specific excess healthcare costs in 2013 Australian dollars.

| **Cancer Type** | **Stage I** | **Stage II** | **Stage III** | **Stage IV** | **Stage Unknown** |
| --- | --- | --- | --- | --- | --- |
| Breast Cancer | \$39,433 | \$52,166 | \$76,716 | \$82,395 | \$78,449 |
| Colorectal Cancer | \$40,081 | \$51,727 | \$94,547 | \$93,348 | \$84,174 |
| Lung Cancer | \$22,192 | \$22,192 | \$52,163 | \$61,481 | \$54,973 |
| Melanoma | \$10,875 | \$20,001 | \$23,835 | \$29,725 | \$25,636 |

Finally, since the costs presented by Goldsbury et al. were reported in 2013 Australian dollars, the stage-specific costs were indexed to 2020 Australian dollars using the Australian Health Index, which resulted in the final estimates that were used in the analysis (Table A5).

**Table A5.** Approximated stage-specific excess healthcare costs in 2020 Australian dollars.

| **Cancer Type** | **Stage I** | **Stage II** | **Stage III** | **Stage IV** | **Stage Unknown** |
| --- | --- | --- | --- | --- | --- |
| Breast Cancer | \$50,699 | \$67,069 | \$98,632 | \$105,934 | \$100,860 |
| Colorectal Cancer | \$51,531 | \$66,504 | \$121,558 | \$120,015 | \$121,077 |
| Lung Cancer | \$28,532 | \$28,532 | \$67,065 | \$79,045 | \$70,678 |
| Melanoma | \$13,981 | \$25,715 | \$30,644 | \$38,216 | \$32,960 |

